# Hydroxicloroquine for Pre-Exposure Prophylaxis for SARS-COV-2

**DOI:** 10.1101/2020.08.31.20185314

**Authors:** Jaime López de la Iglesia, Naiara Cubelos Fernández, Roi Naveiro Flores, Marina Montoro Gómez, Francisco Javier González de Haro, María Ajenjo González, Estefanía Tobal Vicente, María Lamuedra Gil de Gómez, María Teresa Nuevo Guisado, Isabel Torio Gómez, Ana Peñalver Andrada, Nuria Martínez Cao, Paula González Figaredo, Carlos Robles García, Lidia Anastasia Alvarado Machón, Ángeles Lafont Alcalde, José Cesáreo Naveiro Rilo

**Affiliations:** Doctor en Medicina Familiar y Comunitaria. Jefe de estudios de la Unidad Docente Multidisciplinar de Atención Familiar y Comunitaria de GAP León (España); Licenciada en Medicina. GAP León (España); Instituto de Ciencias Matemáticas. Estadística e investigación operativa. Consejo Superior de Investigación Científica (ICMAT-CSIC); Graduado en Medicina. GAP León (España); Doctora en Medicina. GAP León (España); Doctor en Salud Pública y Medicina Preventiva. Unidad Docente de Medicina Familiar y Comunitaria de León (España)

**Keywords:** Hydroxychloroquine, Pre-exposure prophylaxis, COVID-19, SARS-CoV-2

## Abstract

SARS-CoV-2 infection has a high transmission level. At the present time there is not a specific treatment approved but it is known that, in vitro, chloroquine and hydroxychloroquine can inhibit the coronavirus.

**Objective:** verifying if patients with autoimmune diseases that are on treatment with HCQ have less incidence and severity on COVID-19.

**Material and methods:** this is a retrospective cohort study. The exposed cohort was formed by individuals with autoimmune diseases with HCQ treatment. The control cohort was randomly selected using the Health Card database. To deal with confounding variables and evaluate the effect of HCQ on the incidence and severity of SARS-CoV-2 infection, propensity score matching was used. Risk difference and paired percentage difference between exposed and non-exposed groups was estimated.

**Results:** 919 individuals formed the exposed cohort and 1351 the control cohort. After matching, there were 690 patients on each group. During the time of the study, in the exposed group there were 42 (6.1%) individuals with suspected COVID-19, 12(1.7%) with confirmed COVID-19 and 3(0.4%) were hospitalized. In the control group there were 30(4.3%) individuals with suspected COVID-19, 13(1.9%) with confirmed COVID-19 and 2(0.3%) were hospitalized. The risk difference between each cohort was: 0.017(−0.05-0.04) for suspected COVID-19; -0.014(−0.015-0.012) for confirmed COVID-19 and 0.001(−0.007-0.007) for hospitalized patients. There were not significant differences.

**Conclusion:** there is no difference neither on the incidence nor on the severity of COVID-19 between patients with autoimmune diseases with HCQ treatment and patients that do not take HCQ.

## INTRODUCTION

The big morbi-mortality plus the lack of treatments and specific vaccines against SARS-CoV-2 virus, made the scientific community use drugs that had already been used to fight other diseases. Chloroquine (CQ) and hydroxychloroquine (HCQ) used in vitro^1,2^, have shown effectiveness against other viruses, including the responsible of the previous severe acute respiratory syndrome SARS-CoV coronavirus outbreak. This findings plus the wide experience using these drugs and its low price, made them good candidates to be used for prophylaxis and treatment for COVID-19. In vitro trials in China show that CQ and HCQ inhibit the growth of SARS-CoV-2, showing a good antiviral activity for pre-exposure prophylaxis and treatment. HCQ has a stronger effect as a pre-exposure prophylaxis treatment^3^. Other in vitro trials suggest different dosage for both prophylaxis and treatment for SARS-CoV-2^4^.

Small trials show effectiveness using HCQ both on its own^5^ and in combination with macrolides^6,7^ to treat COVID-19. However, more recent evidence demonstrates the contrary^8,9,10^.

The first trial speculating on a post-exposure prophylactic role of HCQ took place in a hospital in South Korea^11^. There was not a control group. HCQ was given to 211 people (189 patients and 22 hospital workers) with a negative result in the Polymerase Chain Reaction (PCR) diagnose, that had previously been in touch with a COVID-19 patient. After a 14 days quarantine, 97% of the subjects kept a negative PCR.

Some clinical trials^12^ and some publications in the literature^13^ say that patients treated with HCQ were less affected or not affected at all by COVID-19. The potential validity of these findings was used by some politicians to make statements about the benefits of antimalarials in the prophylaxis of SARS-CoV-2^14^. Later publications in the COVID-19 Global Rheumatology Alliance express the opposite^15,16^.

Whilst ongoing clinical trials on the prophylaxis of COVID-19 reveal results, we present this study with the aim of evaluating if patients who are on a chronic treatment with antimalarials have less incidence of infection with SARS-CoV-2 and/or a less severe disease, than patients who do not take antimalarials.

## MATERIALS AND METHODS

**Study type:** retrospective cohort study.

**Data source:** the *exposed cohort* is formed by all the patients on a chronic treatment with HCQ in León (Spain). Mostly due to autoimmune diseases: systemic lupus erythematosus (LES) 43%, rheumatoid arthritis (RA) 28% and other rheumatic diseases 29%. This cohort was elaborated with the CONCILYA database, that holds information about the invoicing of pharmaceutical prescriptions. Patients who had been prescribed with CQ or HCQ during December 2019, January and February 2020 were selected using the Patient Identification Number (PIN). We assume that considering this range of time, we selected all the patients that take antimalarials on a chronic base. Data was firstly preprocessed, removing 37 patients who had only withdrawn one package of either CQ or HCQ from the pharmacy in March, verifying that they were not diagnosed with a rheumatic disease. We suppose they had withdrawn it as a possible prevention for COVID-19. Another 42 patients were removed as they did not belong to the study area.

The *non-exposed cohort* was selected randomly sampling from the Individual Health Card (IHC) database of the population from the target area. They were matched by sex, due to the difference in the prevalence of rheumatic diseases between men and women, and by a five-years age range.

**Sample size and power of the study:** the size of the exposed cohort, 919, is given by the number of patients with autoimmune diseases (LES and RA mostly) that take CQ or HCQ. 1351 subjects of the non-exposed cohort were selected in order to get a 95% confidence. In 900 pairs of subjects we would detect risk differences of 95% or more with an 80% power.

### Study variables

*Exposure variables:* taking or not CQ or HCQ. It was classified as a dichotomous variable (yes/no).

*Output variables:* having COVID-19 disease during the months of maximum impact of the pandemic (March and April 2020). This variable was studied in two different levels: possible disease or confirmed disease. Possible corresponds to patients with COVID-19-like symptoms such as non-severe acute respiratory infection (fever, cough, dyspnea, myalgia, shivers), ageusia or anosmia, during the time of the study, with no diagnostic tests. Confirmed, corresponds to having a positive Polymerase Chain Reaction (PCR) test for SARS-CoV-2 in a nasopharyngeal swab or having COVID-19-like symptoms and a positive COVID-19 Rapid Antibody Test (IgM and/or IgG). Being hospitalized was also evaluated to measure the severity of the disease.

*Other variables:* sociodemographic data: age, sex, rural or urban area, sanitary professional, workers at elderly homes; disease for which they take antimalarials; coagulopathy; diseases and drugs that could affect on the morbimortality of COVID-19: obesity, smoker, hypertension (HT), lung disease, cardiovascular disease (CVD); patients on treatment with: corticosteroids, angiotensin-converting-enzyme inhibitors (ACE inhibitor) / angiotensin II receptor blockers (ARBs), anticoagulant medication, vitamin D, non-steroidal anti-inflammatory (NSAIDs).

*Fieldwork:* using the PIN, we had access to each patient personal history database in MEDORA (used in Primary Care) and JIMENA (used in the hospital). It was checked on every patient if they had suffered COVID-19, which symptoms they had and if they had been hospitalized. Each patient was contacted via telephone to obtain variables that were not present on their personal history, such as previous exposure to the virus. May 1st 2020 was considered the study closure date.

### Statistical analysis

A descriptive study of each variable was carried out using frequency distributions for qualitative variables, means and standard deviations for quantitative ones.

This is an observational study. Thus, in order to estimate causal effects of treatment with CQ or HCQ on the different outcomes, it is necessary to control for confounding. For this purpose we use propensity score mating (PSM)^17^. PSM first estimates the propensity score (i.e. the conditional probability of receiving treatment given the confounding variables) for all individuals. Subsequently, individuals of both groups (treatment and control) are matched by propensity scores. As PSM cannot deal with missing data, an imputation process was previously carried out using multiple imputation by chained equations (MICE)^18^. A sensitivity analysis of the conclusions of the study against different imputations of missing values is presented in the Appendix.

The propensity score was estimated using a logistic regression model. The confounding variables included in the model were those that, based on the results of the raw analyses and the bibliography consulted^19^, could behave as confounding factors. Specifically, they were following ones: age, sex, smoker, HT, Diabetes, CVD, Lung disease, anticoagulant medication, corticosteroids, vitamin D, NSAIDs, and possible exposure to COVID. A one-to-three matching was performed using “closest neighbors” technique^20^. A caliper of 0.2 was used (that is, pairs whose distance between propensity scores is greater than 0.2 standard deviations are not accepted). After matching, to ensure balance, differences between treatment and control groups in the all the covariates are checked using standardized mean deviations (SMDs). In the appendix, the conclusions of the study are shown for one-to-two and one-to-four matching.

Finally, after performing PSM, causal effects are estimated in the paired sample. To that end, the risk difference between treatment and control groups was studied for all three outcome variables (using a paired Student test), as well as the difference in paired percentages (through a McNemar test).

All the statistical calculations of the present study were performed using the R statistical software, version 3.6.3. The imputation of missing values was made with the R^21^ “mice” library and the propensity score “matching” with this same software^22^.

## RESULTS

### Descriptive analysis and PSM matching

There were 919 patients with rheumatic diseases on treatment with antimalarials. 55% took 400mg of HCQ per day; 44%, 200mg or less and only 1% took CQ. 85% were on treatment for more than one year. 4% of the patients in the non-treated group and 5.4% of those in the treatment group were diagnosed with possible COVID-19. On 1.6% of each group the diagnosis was confirmed. Table 1 shows the characteristics of the treatment and control groups, before and after matching.

**Table 1.** Individuals characteristics depending on whether if they took HCQ before or after the matching with PSM.

|  | Original sample |  |  | Matching sample after PSM |  |  |
| --- | --- | --- | --- | --- | --- | --- |
|  | Not treated with HCQ | Treated with HCQ | SMD | Not treated with HCQ | Treated with HCQ | SMD |
| **n** | 1351 | 919 |  | 690 | 690 |  |
| **Age (mean(SD))** | 58.96 (15.91) | 57.28 (15.03) | 0,109 | 55.66 (16.76) | 57.40 (14.19) | 0,112 |
| **Women, n (%)** | 1055 (78.1) | 706 (76.8) | 0,03 | 530 (76.8) | 522 (75.7) | 0,027 |
| **Profession, n (%)** |  |  | 0,049 |  |  |  |
| **Healthcare professional** | 23 (1.7) | 21 (2.3) |  |  |  |  |
| **Elderly homes** | 33 (2.4) | 26 (2.8) |  |  |  |  |
| **Others** | 1295 (95.9) | 872 (94.9) |  |  |  |  |
| **Smoker, n (%)** | 227 (16.8) | 164 (17.8) | 0,028 | 130 (18.8) | 147 (21.3) | 0,062 |
| **Urban area, n (%)** | 756 (56.0) | 499 (54.3) | 0,033 |  |  |  |
| **Obesity, n (%)** | 251 (18.6) | 209 (22.7) | 0,103 | 130 (18.8) | 144 (20.9) | 0,051 |
| **HT, n (%)** | 369 (27.3) | 322 (35.0) | 0,167 | 198 (28.7) | 213 (30.9) | 0,048 |
| **Diabetes, n (%)** | 130 (9.6) | 93 (10.1) | 0,017 | 55 (8.0) | 63 (9.1) | 0,041 |
| **CVD, n (%)** | 64 (4.7) | 100 (10.9) | 0,231 | 43 (6.2) | 63 (9.1) | 0,109 |
| **Lung disease, n (%)** | 24 (1.8) | 39 (4.2) | 0,145 | 16 (2.3) | 24 (3.5) | 0,069 |
| **Take ACE inhibitors, n (%)** | 143 (10.6) | 99 (10.8) | 0,006 |  |  |  |
| **Take ARBs, n (%)** | 144 (10.7) | 139 (15.1) | 0,134 |  |  |  |
| **Take anticoagulants, n (%)** | 58 (4.3) | 112 (12.2) | 0,29 | 37 (5.4) | 45 (6.5) | 0,049 |
| **Take corticosteroids n (%)** | 27 (2.0) | 346 (37.6) | 1 | 25 (3.6) | 39 (5.7) | 0,097 |
| **Take vitamin D, n (%)** | 144 (10.7) | 644 (70.1) | 1,522 | 140 (20.3) | 138 (20.0) | 0,007 |
| **Take NSAIDs, n (%)** | 113 (8.4) | 234 (25.5) | 0,468 | 100 (14.5) | 108 (15.7) | 0,032 |
| **Drink tonic, n (%)** | 22 (1.6) | 46 (5.0) | 0,189 |  |  |  |
| **Work COVID-19 exposure, n (%)** | 50 (3.7) | 52 (5.7) | 0,093 | 36 (5.2) | 30 (4.3) | 0,041 |
| **Familiar COVID-19 exposure, n (%)** | 43 (3.2) | 26 (2.8) | 0,021 | 19 (2.8) | 30 (4.3) | 0,086 |
| **Suspected COVID-19, n (%)** | 54 (4) | 50 (5,4) | 0,021 | 30 (4.3) | 42 (6.1) | 0,078 |
| **Confirmed COVID-19, n (%)** | 22 (1.6) | 15 (1.6) | <0.001 | 13 (1.9) | 12 (1.7) | 0,011 |
| **Hospitalized for COVID-19, n (%)** | 5 (0.4) * | 6 (0.7) | 0.04 | 2 (0.3)* | 3 (0.4) | 0,024 |

SD: standard deviation; SMD: standardized mean deviation. HT: hypertension, ACE inhibitors: angiotensin-converting-enzyme inhibitors, ARBs: angiotensine II receptor blockets, NSAIDs: non-steroidal anti-inflammatory.
\*: one patient passed on each group.

As shown by the SMD, before PSM there were great differences between exposed and control groups in covariables such as HT, CVD, lung disease, treatment with corticosteroids, vitamin D, anticoagulants or NSAIDs. This was expectable, as patients with autoimmune diseases (who were on treatment with antimalarials) were more likely to suffer HT or lung diseases. In addition, there is also a strong correlation between taking antimalarials and vitamin D or corticosteroids since this drugs are also part of the treatment for autoimmune diseases.

These variables may act as confounders. To achieve an adequate balance in treatment and control groups and be able to extract causal conclusions we use PSM. As shown in Table 1, after PSM, 690 patients survive within each cohort. Now, the differences in the covariates between the two groups are much smaller, as shown by the SMD. Thus, the matched sample can be considered to be balanced.

### Causal effects estimation

The following results were obtained using matched data: in the exposed cohort there were 42 (6.1%) individuals with suspected COVID-19; 12 (1.7%) individuals with confirmed COVID-19 and 3 (0.4%) individuals were hospitalized. In the non-exposed cohort there were 30 (4.3%) individuals with suspected COVID-19; 13 (1.9%) individuals with confirmed COVID-19 and 2 (0.3%) individuals were hospitalized.

The risk difference between each cohort, as shown in Table 2, was: 0.017 (−0.05-0.04) for suspected COVID-19; -0.014 (−0.015-0.012) for confirmed COVID-19 and 0.001 (−0.007-0.007) for hospitalized patients. There were not significant differences between each cohort on neither of the three variables.

**Table 2.** HCQ effect on having suspected COVID-19, confirmed COVID-19 and being hospitalized for COVID-19, after PSM.

|  | **Suspected COVID** |  | **Confirmed COVID** |  | **Hospitalized for COVID** |  |
| --- | --- | --- | --- | --- | --- | --- |
|  | Yes | No | Yes | No | Yes | No |
| **Treatment with HCQ** | 42 | 648 | 12 | 678 | 3 | 687 |
| **Not treatment with HCQ** | 30 | 660 | 13 | 677 | 2 | 688 |
| **Risk difference (I.C. 95%)** | 0.017<br>(-0.005;0.04) |  | -0.0014<br>(-0.015;0.012) |  | 0,001<br>(-0.007;0.007) |  |
| **McNemar's chi square** | 1.8;<br>p-value=0.17 |  | 0.001;<br>p-value=0,662 |  | 0.00;<br>p-value=1 |  |

## DISCUSSION

Our results show that being on a chronic treatment with either CQ or HCQ has neither benefit on the pre-exposure prophylaxis for SARS-CoV-2 nor on the avoidance of being hospitalized, as a subrogate variable for severity of the infection.

Age, sex, rural or urban area and being an active smoker were balanced on the original sample in both groups. On the contrary, obesity, CVD, HT and lung disease were more frequent on the group formed by individuals who were taking HCQ due to a rheumatic disease. These illness association with autoimmune diseases^23^ could have affected the final results^24^. These same comorbidities plus advanced age, male sex and diabetes were associated with a worse outcome on the SARS-CoV-2 infection^25,26^. A great number of individuals on the exposed cohort were on treatment with corticosteroids, immunosuppressive drugs or biological therapies. There is a controversy in the conclusions of different studies about COVID-19 severity in patients with autoimmune diseases who are on treatment with this kind of drugs^27-30^. All these facts could make us believe that the results of our trial would show a protective effect from the antimalarial drugs. However, after matching, there is no significative difference between both groups neither in the number of hospitalized patients nor in the mortality of COVID-19.

Our results are in line with other studies that reveal a lack of evidence on the efficiency of HCQ for prophylaxis and treatment for COVID-19^31^. A meta-analysis shows poor evidence on the efficiency of CQ and HCQ on the prevention of COVID-19^32^. Likewise, Boulmare DR et al on a clinical trial in the USA and Canada^33^, conclude that HCQ shows no efficiency on the post-exposure prevention neither for suspected nor for confirmed COVID-19. In a Spanish trial on autoimmune inflammatory diseases, Macias J et al^34^ reveal a similar result. They also observe no differences with placebo neither on the treatment for suspected or confirmed COVID-19 nor on the hospitalized individuals. 400mg of HCQ per day was the dosage used by most of our individuals. The same dosage is being used in other ongoing trials for pre-exposure and post-exposure prophylaxis of COVID-19, shown in clinicaltrials.gov (NCT 04333225, NCT 04331834). This dosage is bigger than the one proposed in other in vitro studies^6^ for pre-exposure prophylaxis.

There are several limitations in our study. This is a retrospective study that analyzes real-life data of two groups, treatment and control, whose basal characteristics were not comparable. To overcome this limitation PSM was needed. The size of our sample was reduced because it was not possible to match some individuals.

The estimated number of matches needed to achieve a correct output for the hypothesis tests was 900. Neverthless, after matching, sample size was reduced to 690 patients per group. However, we end up with two comparable groups where the only difference was treatment status before exposure.

In Spain most COVID-19 cases were not severe and therefore were well managed by General Practitioners from Primary Care. At the beginning of the pandemic there were not enough diagnostic tests, hence they were mostly used for hospitalized patients. This is the reason why a considerable number of suspected COVID-19 patients were not confirmed. Of 72 suspected individuals only 25 were confirmed with SARS-CoV-2 infection. The 47 remaining, exposed and non-exposed, were not confirmed. If we would have been able to perform diagnostic tests maybe the confirmed patients would have inclined the balance to either the exposed group or to the non-exposed group.

Having used a pragmatic approach made it easier to recruit the exposed cohort but it made it more difficult to apply these results to general population. All the individuals in the exposed group had rheumatic diseases. This could act as a confounding factor because it presence of rheumatic diseases may be associated with some or the wholesome of the effect variables, this complicating the generalization of the results. Nevertheless, in the hypothetical case where having a rheumatic disease did not affect the outcome variables, the conclusions in this study could be applied to general population. This could confirm the lack of positive benefit of HCQ neither for pre-exposure prophylaxis nor for diminishing severity of COVID-19.

## Data Availability

.

## ACKNOWLEDGMENTS

To León Primary Care Management, for the facilities granted for the study; to the Primary Care Pharmacy Service of the Primary Care Management, for their collaboration in the search of exposed subjects; to Susana González Carrera, for her help in preparing the manuscript; to Rosana Ares, from the Computing Service of the Primary Care Management of León; and to all participants in this study.

RN acknowledgments support of the Spanish Ministry for his grant FPU15-03636.

## CONFLICT OF INTEREST

The authors declare no conflict of interest in this article.

The research protocol was accepted by the Ethics Committee for Drug Research (CEIm) of the Health areas of León and El Bierzo, on April 27, 2020. Internal Registry: 2072.

## COVER LETTER

The following manuscript arises during the Alarm State in our country, because of the universal Pandemic due to the SARS-CoV-2 coronavirus.

Among the drugs that were used as treatment at that time, without any scientific evidence, in the hospital and outpatient settings, was hydroxychloroquine.

For this reason, we asked ourselves the clinical question about whether patients on pre-exposure treatment with Hydroxychloroquine would present a different evolution of the disease, due to the fact that they were previously taking this drug.

## REFERENCES

1. Savarino A, Boelaert JR, Cassone A, Majori G and Cauda R. Effects of chloroquine on viral infections: an old drug against today’s diseases?. Lancet Infect Dis. 2003; 3(11): 722–7

2. Biot C, Daher W, Chavain N, et al. Design and synthesis of hydroxyferroquine derivatives with antimalarial and antiviral activities. J Med Chem. 2006; 4;49(9):2845-9; doi: 10.1021/jm0601856

3. YaoX, YeF, Zhang M, Cui C, Huang B, Niu P. In vitro antiviral activity and projection of optimized dosing design of hydroxychloroquine for the treatment of severe acute respiratory syndrome main point: hydroxychloroquine was found to be more potent than chloroquine at inhibiting SARS-CoV-2 in vit. Clin Infect Dis. 2020; 2:1-25

4. Al-Kofaji M, Jabobson P, Boulware DR, Matas A, Kandaswamy R, M Jaber M, et al. Finding the Dos for Hydroxychloroquine Prophylaxis for COVID‐19: The Desperate Search for Effectiveness. Clinical Pharmacology & Therapeutics. 2020; doi: 10.1002/cpt.1874

5. Zhaowei Chen, Jijia Hu, Zongwei Zhang, Shan Jiang, Shoumeng Han, Dandan Yan, et al. Efficacy of hydroxychloroquine in patients with COVID-19: results of a randomized clinical trial. [Preprint]. March 2020. doi: 10.1101/2020.03.22.20040758. Available from: https://pubmed.ncbi.nlm.nih.gov/16640347/

6. Gautret P, Lagier JC, Parola P, Hoang VT, Meddeb L, Mailhe M et al. Hydroxychloroquine and azithromycin as a treatment of COVID-19: results of an open-label non-randomized clinical trial [Preprint]. March 2020. Int J Antimicrob Agents. 2020;105949. doi:10.1016/j.ijantimicag.2020.105949

7. Million M, Lagier JC, Gautret P, Colson P, Fournier P, Amrane S. Early treatment of COVID-19 patients with hydroxychloroquine and azithromycin: A retrospective analysis of 1061 cases in Marseille, France [Preprint]. May 2020. Travel Med Infect Dis. 2020; 101738. https://doi.org.10.1016/j.tmaid.2020.101

8. Tang W, Cao Z, Han M, Wang Z,Chen J, Sun W. Hydroxychloroquine in patients with mainly mild to moderate coronavirus disease 2019: open label, randomised controlled trial. BMJ May 2020; 369 doi: https://doi.org/10.1136/bmj.m1849

9. Mahévas M, Tran VT, Roumier M, Chabrol A, Paule R, Guillaud C. Clinical efficacy of hydroxychloroquine in patients with COVID-19 pneumonia who require oxygen: observational comparative study using routine care data. BMJ. 2020; 369. Doi: https://doi.org/10.1136/bmj.m1844

10. Rosenberg ES, Dufort EM, Udo T, Wilberschied LA, Kumar J, Tesoriero J, et al. Association of Treatment With Hydroxychloroquine or Azithromycin With In-Hospital Mortality in Patients With COVID-19 in New York State. JAMA 2020; 1-10. Doi: 10.1001/jama.2020.8630

11. Lee S, Son H, Ran Peck K. Can post-exposure prophylaxis for COVID-19 be considered as an outbreak response strategy in long-term care hospitals? Int J Antimicrob Agents. April 2020. doi:10.1016/j.ijantimicag.2020.105988.URL:https://www.ncbi.nlm.nih.gov/pmc/articles/PMC7162746/

12. Chen Z, Hu J, Zhang Z, Jiang S, Han S, Yan D. Efficacy of hydroxychloroquine in patients with COVID-19: results of a randomized clinical trial. [Preprint]. Epidemiology 2020. doi:10.1101/2020.03.22.20040758

13. Joob B, Wiwanitkit V. SLE, hydroxychloroquine and no SLE patients with COVID-19: a comment. Ann Rheum Dis. 2020;79(6):e61. doi:10.1136/annrheumdis-2020-217506. Available from: http://www.ncbi.nlm.nih.gov/pubmed/32295787

14. James S.Brady Press Briefing Room. Remarks by President Trump, Vice President Pence, and Members of the Coronavirus Task Force in Press Briefing. Available from: https://www.whitehouse.gov/briefings-statements/remarks-president-trump-vice-president-pence-members-coronavirus-task-force-press-briefing-31/

15. Mathian A, Mahevas M, Rohmer J, Roumier M, Cohen-Aubart, Amador-Borrego B. Clinical course of coronavirus disease 2019 (COVID-19) in a series of 17 patients with systemic lupus erythematosus under long-term treatment with hydroxychloroquine. Ann Rheum Dis. 2020;79 (6):837‐839. doi:10.1136/annrheumdis-2020-217566

16. Konig MF, Kim AH, Scheetz MH,Graef ER, Liew JW, Simard J. Baseline use of hydroxychloroquine in systemic lupus erythematosus does not preclude SARS-CoV-2 infection and severe COVID-19. Annals of the Rheumatic Diseases. Published Online First: 07 May 2020. doi: 10.1136/annrheumdis-2020-217690

17. Rosenbaum, P. R., & Rubin, D. B. The central role of the propensity score in observational studies for causal effects. Biometrika. 1983. 70(1), 41-55.

18. Buuren, S. V., & Groothuis-Oudshoorn, K. Mice: Multivariate imputation by chained equations in R. Journal of statistical software 2010, 1-68.

19. Emmi G, Bettiol A, Mattioli I, et al. SARS-CoV-2 infection among patients with systemic autoimmune diseases. Autoimmun Rev. 2020; 19(7):102575. doi:10.1016/j.autrev.2020.102575)

20. Becker S, Ichino A. Estimation of average treatment effects based on propensity scores. The Stata journal. 2002. 2: 358-77

21. Buuren, S. V., & Groothuis-Oudshoorn, K. Mice: Multivariate imputation by chained equations in R. Journal of statistical software. 2010. 1-68

22. Sekhon, J. S. (2008). Multivariate and propensity score matching software with automated balance optimization: the matching package for R. Journal of Statistical Software. 2011. 42(7). Doi: http://dx.doi.org/10.18637/jss.v042.i07

23. Villa A, Mandell BF. Trastornos cardiovasculares y enfermedad reumatica. Rev Esp Cardiol. 2011; 64(9): 809–817

24. Mehra MR, Desai SS, kuy S, henry TD, Patel TD. Cardiovascular Disease, Drug Therapy, and Mortality in COVID-19 (published on line a head of print). N Engl J Med. May 2020; NEJMoa2007621.doi: 10.1056/NEJMoa2007621

25. Cummings MJ, Baldwin MR, Abrams D, et al. Epidemiology, clinical course, and outcomes of critically ill adults with COVID-19 in New York City: a prospective cohort study [published online ahead of print, 2020 May 19]. Lancet. 2020. S0140-6736(20)31189-2

26. Zhou F, Yu T, Du R, et al. Clinical course and risk factors for mortality of adult In patients with COVID-19 in Wuhan, China: a retrospective cohort study. Lancet. 2020. 395(10229):1054‐1062

27. Pablos JL, Abasolo-Alcázar L, Álvaro-Gracia JM, Blanco FJ et al. Prevalence of hospital PCR confirmed COVID-19 cases in patients with chronic inflammatory and autoimmune rheumatic diseases. Ann Rheum Dis, 2020. Doi: https://doi.org/10.1101/2020.05.11.20097808

28. D’Silva KM, Serling-Boyd N, Wallwork R, et al. Clinical characteristics and outcomes of patients with coronavirus disease 2019 (COVID-19) and rheumatic disease: a comparative cohort study from a US ‘hot spot’ [published online ahead of print, 2020 May 26]. Ann Rheum Dis. 2020;doi: https://doi.org/10.1136/annrheumdis-2020-217888

29. Gianfrancesco M, Hyrich KL, Al-Adely S, et al. Characteristics associated with hospitalisation for COVID-19 in people with rheumatic disease: data from the COVID 19 Global Rheumatology Alliance physician-reported registry [published online ahead of print, 2020 May 29]. Ann Rheum Dis. 2020.

30. Pablos JL, Galindo M, Carmona L, Retuerto M, Lledó A, Blanco R, González-Gay MA, et al. Clinical Outcomes of Patients with COVID-19 and Chronic Inflammatory and Autoimmune Rheumatic Diseases: A Multicentric Matched-Cohort Study. [Preprintt].2020. doi: https://doi.org/10.1101/2020.06.18.20133645

31. Shah S, Das S, Jain A, Misra DP, Negi VS. A systematic review of the prophylactic role of chloroquine and hydroxychloroquine in coronavirus disease-19 (COVID-19). Int J Rheum Dis. 2020;23(5):613-619. doi:10.1111/1756-185X.13842)

32. Cavalcanti AB, Zampieri FG, Regis R, et al. Hydroxychloroquine with or without Azithromycin in Mild-to-Moderate Covid-19. NEJM. June 2020. Doi: 10.1056/NEJMoa2019014

33. Boulware DR, Pullen MF, Bangdiwala AS, et al. A randomized trial of hydroxychloroquine as postexposure prophylaxis for COVID-19. N Engl J Med. June 2020. doi: 10.1056/NEJMoa2016638.

34. Macias J, Gonzalez-Moreno P, Sanchez-Garcia E, Morillo-Verdugo R, Dominguez-Quesada C, Pinilla A. Similar incidence of Coronavirus Disease 2019 (COVID-19) in patients with rheumatic diseases with and without hydroxychloroquine therapy. [Preprint] 2020. Doi: https://doi.org/10.1101/2020.05.16.20104141

